# Bounding the Predictive Values of COVID-19 Antibody Tests

**DOI:** 10.1101/2020.05.14.20102061

**Authors:** Charles F. Manski

## Abstract

COVID-19 antibody tests have imperfect accuracy. There has been lack of clarity on the meaning of reported rates of *false positives* and *false negatives*. For risk assessment and clinical decision making, the rates of interest are the positive and negative predictive values of a test. *Positive predictive value* (PPV) is the chance that a person who tests positive has been infected. Negative predictive value (NPV) is the chance that someone who tests negative has not been infected. The medical literature regularly reports different statistics, *sensitivity* and *specificity*. Sensitivity is the chance that an infected person receives a positive test result. Specificity is the chance that a non-infected person receives a negative result. Knowledge of sensitivity and specificity permits one to predict the test result given a person’s true infection status. These predictions are not directly relevant to risk assessment or clinical decisions, where one knows a test result and wants to predict whether a person has been infected. Given estimates of sensitivity and specificity, PPV and NPV can be derived if one knows the *prevalence* of the disease, the rate of illness in the population. There is considerable uncertainty about the prevalence of COVID-19. This paper addresses the problem of inference on the PPV and NPV of COVID-19 antibody tests given estimates of sensitivity and specificity and credible bounds on prevalence. I explain the methodological problem, show how to estimate bounds on PPV and NPV, and apply the findings to some tests authorized by the FDA.

I am grateful to Michael Gmeiner for helpful comments.

## 1. Introduction

There are now two main classes of tests for COVID-19. Nasal swab tests detect the presence of live virus within a person, signaling an active infection. Serological tests detect the presence of antibodies that the immune system develops after onset of infection. The presence of antibodies signals that a person was infected at some past date.

Tests of both types have imperfect accuracy. A *false positive* occurs when a test result indicates infection, but the person has not actually been infected. A *false negative* occurs when a result indicates no infection, but the person has been infected. These concepts are transparent. Yet there has been lack of clarity on the meaning of reported rates of false positives and negatives. It is important to interpret them correctly.

For personal risk assessment and clinical decision making, the rates of interest are the positive and negative predictive values of a test. *Positive predictive value* (PPV) is the chance that a person who tests positive has indeed been infected. Negative predictive value (NPV) is the chance that someone who tests negative has not been infected.

The medical literature on test accuracy regularly reports statistics other than PPV and NPV, known as *sensitivity* and *specificity*. Sensitivity is the chance that an infected person receives a positive test result. Specificity is the chance that a non-infected person receives a negative result.

Knowledge of sensitivity and specificity permits one to predict the test result given a person’s true infection status. These predictions are not directly relevant to risk assessment or clinical decisions. For these purposes, one knows a test result and wants to predict whether a person has been infected. One does not know whether a person has been infected and want to predict a test result.

Perceptive writers on diagnostic testing have long warned that PPV-NPV are relevant to risk assessment and clinical decisions, whereas sensitivity and specificity are not. Close to thirty years ago, Altman and Bland (1994) wrote:

> “The whole point of a diagnostic test is to use it to make a diagnosis, so we need to know the probability that the test will give the correct diagnosis. The sensitivity and specificity do not give us this information. Instead we must approach the data from the direction of the test results, using predictive values. Positive predictive value is the proportion of patients with positive test results who are correctly diagnosed. Negative predictive value is the proportion of patients with negative test results who are correctly diagnosed.”

The warning expressed by Altman and Bland and other writers should be heeded.

Given that PPV-NPV are clinically relevant concepts while sensitivity and specificity are not, it is natural to ask why the recent medical research on testing for COVID-19 focuses mainly on the latter quantities rather than the former. In the case of antibody tests, part of the answer appears to be that medical researchers find it easier to measure sensitivity and specificity.

Estimation of PPV-NPV requires accurate observation of test results and true infection status for a random sample of the relevant population. It is easy to observe rest results but not true infection status. If it were easy to observe true infection status, diagnostic tests would serve no practical purpose.

Whereas observation of true infection status for a random sample of persons is difficult, researchers have found it possible to do so using data from special samples of persons whose infection status is known. This enables estimation of sensitivity and specificity, at least for these special samples. The practice has been to estimate in this manner and to assume that the findings obtained for the special samples hold more generally.

Consider specificity. The virus being novel, it is known that antibodies cannot be present in blood drawn from humans prior to the late 2019 onset of the pandemic. Blood drawn from numerous persons is held in storage in various locations for multiple purposes. Applying a serological test for antibodies to samples of this stored blood should always yield a negative result. Positive test results must be false positives. Thus, the specificity of a COVID-19 antibody test when administered to blood drawn prior to late 2019 can be estimated by applying the test to samples of such blood. The U.S. Food and Drug Administration (FDA) states the idea as follows (FDA, 2020): “A test’s specificity can be estimated by testing large numbers of samples collected and frozen before SARS-CoV-2 is known to have circulated to demonstrate that the test does not produce positive results in response to the presence of other causes of a respiratory infection, such as other coronaviruses.”

Consider sensitivity. Clinicians are confident that they can determine that persons have been infected using a highly sensitive type of test thought to have perfect accuracy. FDA (2020) states: “A test’s sensitivity can be estimated by determining whether or not it is able to detect antibodies in blood samples from patients who have been confirmed to have COVID-19 with a nucleic acid amplification test, or NAAT.” Applying a serological test for antibodies to samples of blood from patients who have tested positive by the NAAT method should always yield a positive result. Negative test results must be false negatives. Thus, the sensitivity of a COVID-19 antibody test when administered to blood drawn from patients who have been subjected an NAAT test can be estimated by applying the test to samples of such blood.

Suppose that sensitivity and specificity have been estimated as described above and that one finds it credible to extrapolate the findings from the special samples to the relevant patient population. PPV-NPV can then be derived if one knows the *prevalence* of the disease; that is, the marginal rate of illness in the population. In the case of COVID-19, prevalence is the population infection rate. To derive estimates of PPV-NPV, FDA (2020) assumes that the infection rate is 0.05. However, the FDA recognizes that this assumption lacks foundation, stating (original in bold): “**We do not currently know the prevalence of SARS-CoV-2 antibody positive individuals in the U.S. population, and prevalence may change based on the duration the virus is in the country and the effectiveness of mitigations.”**

A primary source of uncertainty about the COVID-19 infection rate is a serious problem of missing data. Confirmed cases have commonly been measured by rates of positive nasal swab findings among persons who have been tested for infection. Infection data are missing for persons who have not been tested. The persons who have been tested differ considerably from those who have not been tested. Criteria used to determine who is eligible for testing typically require demonstration of symptoms associated with presence of infection or close contact with infected persons. This gives considerable reason to believe that some fraction of untested persons are asymptomatic or pre-symptomatic carriers of the COVID-19 disease. Presuming this is correct, the actual cumulative rate of infection has been higher than the reported rate.

Manski and Molinari (2020) analyze the inferential problem and report credible location and time-specific bounds on the infection rate. The bounds are wide. For example, on April 24, 2020, the bounds on the infection rates in Illinois and New York were [0.004, 0.525] and [0.017, 0.618]. Thus, the FDA estimates of PPV-NPV assuming that the infection rate is 0.05 are not well grounded.

This paper addresses the problem of inference on the PPV-NPV of COVID-19 antibody tests given estimates of sensitivity and specificity and credible bounds on prevalence. I explain the methodological problem, show how to estimate bounds on PPV-NPV, and apply the findings to some tests authorized by the FDA.

## 2. Methods

Consider the problem of predicting whether a person in a population of interest has been infected, given observation of an antibody test result. Let y = 1 denote when a person has been infected and y = 0 otherwise. Let x = p denotes a positive test result and x = n a negative result. A false positive result occurs when x = p but y = 0. A false negative means that x = n but y = 1.

Let P(y, x) denote the population distribution of illness and test results. The marginal probability P(y = 1) is the prevalence of the illness and P(x = p) is the rate of positive test results. Assume that 0 < P(y = 1) < 1 and 0 < P(x = p) < 1. Then the conditional probability distributions P(y|x) and P(x|y) are well-defined. PPV and NPV are the conditional probabilities P(y = 1|x = p) and P(y = 0|x = n). Sensitivity and specificity are the conditional probabilities P(x = p|y = 1) and P(x = n|y = 0).

If one can observe data on (y, x) randomly drawn from P(y, x), one can estimate PPV-NPV using standard statistical methods. The concern is inference when one does not have random sample data, but one has data that are informative about some aspects of P(y, x). One may have information on test sensitivity P(x = p|y = 1), specificity P(x = n|y = 0), the rate of positive test results P(x = p), and/or the population infection rate P(y = 1).

Suppose that sensitivity and specificity are known. Bayes Theorem and the Law of Total Probability provide the mathematical relationship connecting PPV-NPV with sensitivity and specificity.

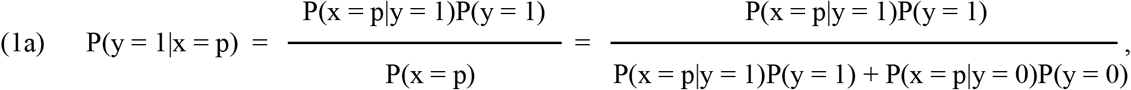

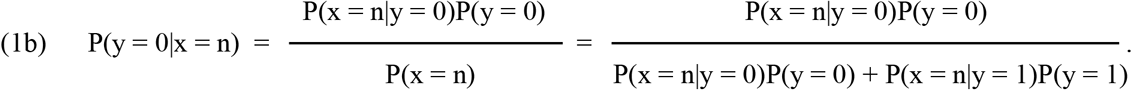

Inspection of the right-hand-side shows that one can compute PPV-NPV if one can combine knowledge of sensitivity and specificity with knowledge of the population infection rate. To estimate PPV-NPV, the FDA combines estimates of sensitivity and specificity with the assumption that the population infection rate P(y = 1) = 0.05. As discussed earlier, this assumption lacks foundation.

Suppose that one has no knowledge of P(y = 1). Then inspection of (1a)-(1b) shows that no conclusions can be drawn on the magnitudes of PPV and NPV if specificity or sensitivity is respectively less than one. As P(y = 1) increases from 0 to 1, equation (1a) shows that P(y = 1|x = p) increases from 0 to 1. As P(y = 0) increases from 0 to 1, (1b) shows that P(y = 0|x = n) increases from 0 to 1.

Now consider an intermediate situation in which one does not know P(y = 1) but one can place credible lower and upper bounds on its value, say P_L_(y = 1) ≤ P(y = 1) ≤ P_U_(y = 1). For example, one might use the bounds derived in Manski and Molinari (2020). Then inspection of (1a)-(1b) shows that the bound on P(y = 1) implies bounds on PPV-NPV.

PPV is an increasing function of P(y = 1), so

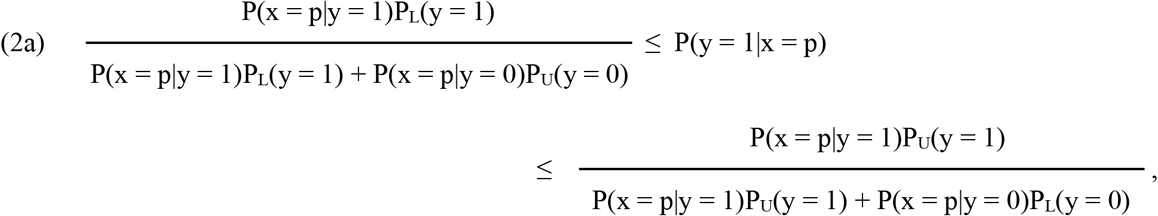

where PU(y = 0) = 1 – P_L_(y = 1) and P_L_(y = 0) = 1 – P_U_(y = 1). NPV increases with P(y = 0), so

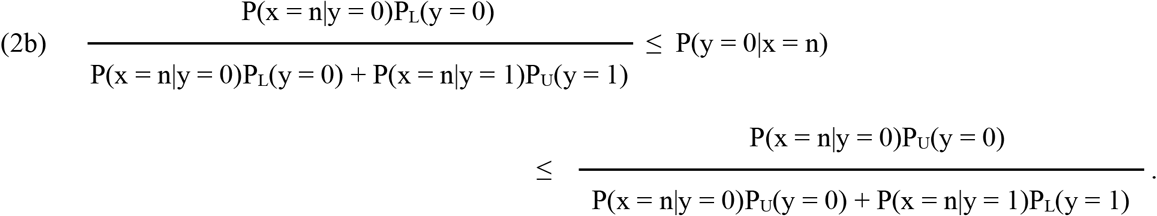

Note the asymmetric manner in which the bound P_L_(y = 1) ≤ P(y = 1) ≤ P_U_(y = 1) on prevalence determines the bounds on PPV and NPV. PPV increases with prevalence, so the lower bound on prevalence yields the lower bound on PPV and the upper bound on prevalence yields the upper bound on PPV. NPV decreases with prevalence, so the upper bound on prevalence yields the lower bound on NPV and the lower bound on prevalence yields the upper bound on NPV. Section 3 will give examples of this asymmetry.

Bounds (2a)-(2b) are a simple application of the type of analysis performed in econometric research on partial identification. See, for example, Manski (2003, 2007). Analysis of partial identification aims to avoid making unjustified strong assumptions about the magnitudes of unknown quantities, such as the COVID-19 infection rate. Instead, the objective is to determine the inferences that are feasible with weaker but more credible assumptions. The results are bounds on quantities of interest rather than precise valuations.

Further results on partial identification of PPV-NPV, beyond those needed in this paper, have been proved in econometric research on inference under *response-based sampling*. In the terminology of that subject, a test result is a covariate and illness status is a response. The research objective is to learn the distribution of response conditional on covariates. The sampling process reveals the distribution of covariates conditional on responses. See Manski (2003, Chapter 6) and Manski (2007, Chapter 6).

## 3. Application to COVID-19 Antibody Tests

FDA (2020) reports estimates of sensitivity and specificity for antibody tests developed by twelve companies, each of which was granted an Emergency Use Authorization. It reports estimates of PPV-NPV under the assumption that the population infection rate equals 0.05. In most cases, the estimates of either or both of sensitivity and specificity are based on blood drawn from a small number of persons. I focus on three tests in which each of the samples used to estimate sensitivity and specificity had at least one hundred observations.

Table 1 uses the sensitivity and specificity estimates in FDA (2020) to estimate bounds on PPV and NPV. The sensitivity and specificity estimates are plugged in for P(x = p|y = 1) and P(x = n|y = 0) in (2a)-(2b). The bounds are computed under the assumption that the prevalence P(y = 1) lies in the interval [0.017, 0.618] reported by Manski and Molinari (2020) for New York on April 24, 2020.

**Table 1:**
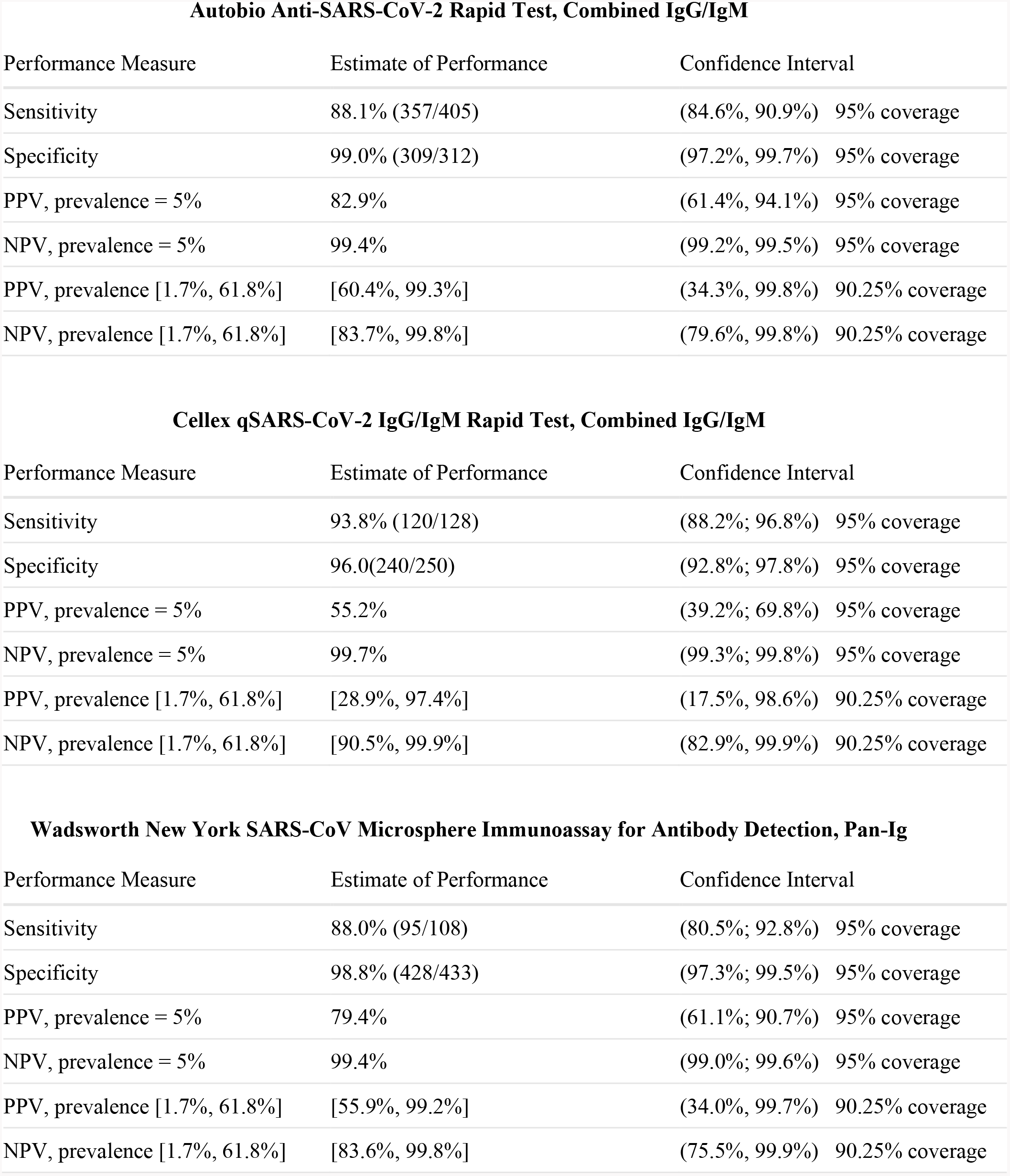
Test Performance.

The table also shows confidence intervals for the partially identified PPV and NPV. The lower (upper) limit of each confidence interval is computed using the lower (upper) limits of the confidence intervals for sensitivity and specificity stated in FDA (2020). The latter confidence intervals have nominal coverage 0.95. Hence, the confidence intervals derived for PPV and NPV have nominal coverage 0.95^2^ = 0.9025. The heading for each panel gives the name and type of the antibody test in which sensitivity and specificity are measured. The first two rows show the sensitivity and specificity estimates reported in FDA (2020). The third and fourth rows show the FDA estimates and confidence intervals for PPV and NPV, assuming P(y = 1) = 0.05. The fifth and sixth rows show the estimated bounds and confidence intervals for PPV and NPV, computed using (2a) and (2b).

For all three tests, the table shows a striking asymmetry in the positions and widths of the estimated bounds for PPV and NPV. The estimated lower bounds on NPV are all greater than 80% and the upper bounds are near 100%. The associated confidence intervals are only slightly wider, with lower bounds greater than 75% and upper bounds again near 100%. Thus, persons receiving negative test results can be reasonably confident that they do not have antibodies to COVID-19.

In contrast, the estimated bounds on PPV have widths ranging from about 40% to 70%, with even wider confidence intervals. The upper bounds are all near 100%. The problem for risk assessment is that the lower bounds are quite low in magnitude, being 60.4%, 28.9%, and 55.9%. The lower bounds of the confidence intervals are considerably lower still. Thus, persons receiving positive test results should not be confident that they have antibodies to COVID-19.

The asymmetry in the bounds on PPV and NPV is a consequence of the asymmetry in how the bound [0.017, 0.618] on prevalence determines these bounds, noted in Section 2. The lower bound on PPV occurs when prevalence is 0.017 and the upper bound when prevalence is 0.618. The lower bound on NPV occurs when prevalence is 0.618 and the upper bound when prevalence is 0.017. The lowest prevalence value 0.017 is much closer to zero than the highest value 0.618 is to one.

## 4. Conclusion

COVID-19 antibody tests have imperfect accuracy. For risk assessment and clinical decision making, test accuracy should be measured by PPV and NPV. Yet the medical literature regularly reports sensitivity and specificity. Given estimates of sensitivity and specificity, PPV and NPV can be derived if one knows the prevalence of the disease, but there is considerable uncertainty about the prevalence of COVID-19. This paper has shown how to derive bounds on PPV and NPV given estimates of sensitivity and specificity and credible bounds on prevalence. Applying these bounds to tests authorized by the FDA, narrow bounds for NPV and wide bounds for PPV are found to hold with the current limited knowledge of prevalence.

## Data Availability

Data referred to in the manuscript were made public by the FDA.

https://www.fda.gov/medical-devices/emergency-situations-medical-devices/eua-authorized-serology-test-performance

## References

Altman, D. and M. Bland (1994), “Statistics Notes: Diagnostic tests 2: predictive values,” BMJ 309, 102.

Manski, C. (2003), Partial Identification of Probability Distributions. New York: Springer-Verlag.

Manski, C. (2007), Identification for Prediction and Decision. Cambridge, MA: Harvard University Press.

Manski, C. and F. Molinari (2020), “Estimating the COVID-19 Infection Rate: Anatomy of an Inference Problem,” Journal of Econometrics, https://doi.org/10.1016/j.jeconom.2020.04.041.

U.S. Food and Drug Administration (2020), EUA Authorized Serology Test Performance, https://www.fda.gov/medical-devices/emergency-situations-medical-devices/eua-authorized-serology-test-performance, accessed May 10, 2020.

